# When Algorithms Prescribe: A Cross-Sectional Study of Quality, Misinformation, and Engagement in Statin-Related Content on TikTok

**DOI:** 10.64898/2026.06.04.26354962

**Authors:** Iren Gharibyan, Emily Ahner, Ryan Shao, Divyansh Sharma, Tracy Navarsartian Tazehkand, Jimmy Diep, Bertille Assoumou

## Abstract

**Background:** Statins are key to preventing atherosclerotic cardiovascular disease and lowering low-density lipoprotein cholesterol and cardiovascular events. However, skepticism regarding their safety and value persists and is increasingly influenced by social media. TikTok has emerged as a major source of health information, but its content varies in quality and accuracy. This study evaluated the quality, attitudes, misinformation, and engagement of statin-related content on TikTok.

**Methods:** Public TikTok videos were collected using predefined search terms and coded by creator type, thematic content, and overall attitude. Video quality was assessed using the DISCERN instrument, the Patient Education Materials Assessment Tool for Audiovisual Materials, and the Global Quality Score. False or misleading claims were independently reviewed by two cardiology fellows. Associations between engagement and quality were also examined.

**Results:** Of 1,349 screened videos, 258 met inclusion criteria. Most were educational (91.0%), with non-physician healthcare providers (34.5%) as the largest creator group. Risks or negative effects were discussed more often than benefits (63.2% vs 42.2%), and 39.5% contained at least one false or misleading claim, most often from complementary and alternative medicine providers and wellness promoters. Quality differed by creator type across all instruments, with physician-created content scoring highest. Video popularity showed minimal association with informational quality.

**Conclusion:** Statin-related TikTok content frequently emphasizes harms, often contains misinformation, and varies substantially in quality by creator type. Greater involvement of healthcare professionals on social media may help improve digital health literacy and counter misleading information about statin therapy.

## 1. Introduction

Cardiovascular disease remains the leading cause of morbidity and mortality worldwide, accounting for an estimated 17.9 million deaths annually [1]. Elevated low-density lipoprotein (LDL) cholesterol is a major causal driver of atherosclerotic cardiovascular disease, as LDL particles infiltrate the arterial wall, promote plaque formation, and increase the risk of atherosclerosis [2]. Statin therapy, also known as hydroxymethylglutaryl-CoA reductase inhibitors, is one of the most effective and widely prescribed interventions for reducing low-density lipoprotein cholesterol and preventing atherosclerotic cardiovascular disease [3,4]. Despite their proven benefits and inclusion in major guidelines, concerns about side effects, overprescription, and mistrust of pharmaceuticals and clinicians continue to fuel skepticism around statin therapy [5–7]. These perceptions contribute to high rates of discontinuation and nonadherence, with direct implications for cardiovascular outcomes [8].

Over the past two decades, social media has become a dominant source of health information, with people of all ages increasingly using these platforms to seek and share medical knowledge [9]. Lim et al. [10] conducted a prospective cohort study of high–cardiovascular-risk patients in Malaysia and found that individuals who actively sought online health information had greater concerns about statins, which were associated with lower medication adherence, suggesting that online content that potentially includes misinformation, may negatively influence treatment behavior.

Other research examining statin-related content on social media has primarily focused on text-based platforms such as Twitter (now X) and Reddit. Golder et al. [11] conducted a qualitative content analysis of 11,852 statin-related Twitter posts, ultimately categorizing 5,201 health-related tweets by user type and topic to identify themes such as beliefs, attitudes, adherence, and adverse events. This study revealed that social media discussions reflect a wide range of perceptions that may influence statin use. Similarly, Hasan et al. [12] analyzed Indonesian Twitter posts about hyperlipidemia medications, finding that most tweets conveyed personal experiences, treatment suggestions, or user perceptions, underscoring social media’s value in assessing public sentiment toward lipid-lowering therapy. Kheloufi et al. [13] examined patient narratives posted to online health forums describing adverse drug reactions (ADRs) to statins and observed that musculoskeletal and nervous system symptoms were most commonly reported, though most posts lacked sufficient clinical detail for pharmacovigilance. Topaz et al. [14] compared adverse drug reactions (ADRs) identified in electronic health records with those discussed on social media, demonstrating that for atorvastatin, the types of side effects patients described online generally mirrored those documented by clinicians. Likewise, Huesch [15] used Facebook data mining to quantify statin-related discussions, reporting that approximately 4% of posts referenced side effects and concluding that social-media monitoring could complement traditional drug-safety surveillance.

More recently, advanced artificial intelligence methods have been applied to characterize public discussions about statins on Reddit. Somani and colleagues [16] analyzed more than 10,000 posts, identifying six thematic groups of discourse, including statin adverse effects, hesitancy, and pharmaceutical industry bias. Their analysis showed that sentiment was predominantly neutral to negative, with only a small minority of discussions portraying statins positively [16]. This work demonstrates the depth and persistence of online skepticism and highlights the power of social media as a lens for understanding public perceptions of therapy.

Platforms such as TikTok have become popular destinations for individuals seeking health advice, yet evidence suggests that inaccurate posts often generate more user engagement than those sharing accurate information [17]. A growing body of studies shows that TikTok can rapidly amplify misinformation across a range of health topics, including reproductive health, sinusitis, and COVID-19 [18–22]. TikTok’s short-form video format and algorithm-driven content distribution amplify material that align with users’ engagement preferences, and its prioritization of engagement over accuracy has been shown to promote the spread of harmful misinformation [23,24].

Although statins play a critical role in cardiovascular disease prevention and online misinformation is well documented, relatively few studies have examined statin-related content on TikTok, and systematic analyses of this content remain limited. Given TikTok’s rapidly expanding influence and its ability to shape health decision-making, a critical gap remains in understanding how this platform influences public perceptions of statin therapy.

Our study addresses this gap by systematically analyzing statin-related TikTok videos, a uniquely algorithm-driven platform where short-form content can rapidly shape engagement and misinformation trends. Using the DISCERN instrument, the Patient Education Materials Assessment Tool for Audiovisual Materials (PEMAT-AV), and the Global Quality Score (GQS), we first assess the quality, accuracy, actionability, and understandability of statin-related TikTok content across creator types, including physicians, wellness influencers, other healthcare professionals, and laypersons. We then characterize each video by thematic content, overall attitude toward statins, and the balance of benefits and risks discussed to better understand how statin therapy is framed on TikTok. Next, we identify the frequency and recurring themes of false or misleading claims and determine which creator groups most frequently share them. Finally, we examine how these content characteristics relate to engagement metrics to provide insight into how statin-related information is communicated and amplified within the TikTok ecosystem.

## 2. Methods

### 2.1. Search Strategy and Data Collection

This study was deemed exempt by the University of Nevada, Las Vegas Institutional Review Board (UNLV-2025-663; approved November 6, 2025), as it involved analysis of publicly available social media content.

Data was collected on November 14, 2025 from publicly available TikTok videos using an automated scraper hosted on the Apify platform. This tool systematically retrieves the top-viewed publicly available videos corresponding to the specified search terms and extracts associated engagement metrics, including views, likes, comments, shares, and saves. As the scraper operates automatically on publicly accessible content, no new TikTok accounts, including accounts without prior viewing history, were created for the search process. Because the search process was automated, the initial search and scraper execution were performed by one researcher. Engagement data obtained through the scraper were subsequently verified by two video reviewers.

Search terms were developed based on the most commonly prescribed statins in the United States. Large-scale analyses demonstrate that atorvastatin and simvastatin have historically been the most frequently prescribed statins in the United States, with simvastatin (41.4%) and atorvastatin (28.3%) representing the majority of use in 2012–2013, followed by pravastatin, rosuvastatin, and lovastatin (Salami, 2017). More recent data indicate that in 2019, atorvastatin remained the most prescribed statin, followed by simvastatin, rosuvastatin, pravastatin, and lovastatin (Matyori, 2023). Brand names were also included due to high public recognition and historical market prominence. For example, Zocor (simvastatin) and Lipitor (atorvastatin) were among the top-selling medications globally prior to loss of market exclusivity [27]. Because TikTok’s search algorithm does not publicly disclose whether singular and plural terms are automatically grouped, both forms were included to maximize capture [28]. The final search terms were: statin, statins, statin medication, cholesterol medication, cholesterol pills, atorvastatin, rosuvastatin, simvastatin, pravastatin, lovastatin, Lipitor (atorvastatin), Crestor (rosuvastatin), Zocor (simvastatin), Pravachol (pravastatin), Mevacor (lovastatin).

### 2.2. Inclusion and Exclusion Criteria

The scraper yielded 1,349 unique videos posted from public TikTok accounts. Videos were eligible for inclusion if they (1) were publicly accessible at the time of extraction, (2) were presented in English, and (3) covered topics relevant to statin medications. Videos were excluded if they were non-English or if their content was unrelated to statins upon manual review. Two authors independently screened all videos for eligibility. Discrepancies were resolved through discussion with a third author. Videos were also excluded if they were deleted or became unavailable prior to the completion of analysis.

### 2.3. Data Extraction and Coding

#### 2.3.1. Video metrics

The TikTok scraper used to collect the publicly available videos automatically collected video length and engagement metrics. Extracted engagement variables included total views, likes, comments, shares, and saves. Engagement rate was calculated as the sum of likes, comments, shares, and saves divided by total views.

#### 2.3.2. Creator qualifications and video categories

Reviewers coded creator qualifications and video category using predefined operational definitions. Creators were categorized as physicians (MD or DO credentials); non-physician healthcare providers (e.g., nurse practitioner, physician assistant, nurse, medical assistant, certified nursing assistant, pharmacist, pharmacy technician); complementary and alternative medicine (CAM) professionals (e.g., naturopathic doctor, chiropractor, therapist, physical therapist, occupational therapist); wellness promoters including individuals identifying as lifestyle coaches, fitness influencers, or holistic advocates or laypersons without healthcare credentials.

Videos were categorized according to their primary intent as educational, anecdotal, entertainment, or other. These video intent and content categories were defined based on categorization approaches used in similar social media health content analyses in the literature [20,22,29]. Educational videos were defined as those primarily intended to inform or teach viewers about a health-related topic, regardless of the accuracy of the information presented. Anecdotal videos featured personal experiences or individual accounts related to statin use. Entertainment videos were created primarily to amuse or engage viewers through humor, satire, or trends rather than to educate or share personal experience. Videos that did not clearly fit into one of these categories were classified as other. Reviewers also categorized each video based on the topic(s) discussed. Topics included discussion of statin mechanism of action; benefits or positive effects; risks or negative effects; misinformation or controversy surrounding statin use; alternatives to statin therapy; public health and advocacy messaging; and lifestyle modifications.

Additional data collected regarding the content of the videos were positive and negative effects of statin therapy as described by the creator, regardless of whether the statements were medically accurate. These categories were selected based on clinically recognized and commonly discussed effects of statins that were expected to appear frequently in social media discourse. Positive effects were informed by the established benefits of statins in lowering low-density lipoprotein cholesterol and reducing cardiovascular events, and negative effects were based on the adverse effects most commonly associated with statin use in the clinical literature and statin FDA drug labels [31–35]. Positive effects were further categorized into heart health, stroke prevention, longevity, cholesterol lowering, safety or minimal side effects, and other. Negative effects were categorized as muscle pain or weakness, liver damage, cognitive changes (including memory loss, brain fog, confusion, or difficulty concentrating), gastrointestinal effects (nausea, vomiting, abdominal pain, diarrhea, constipation, bloating), fatigue, metabolic or diabetes risk, skin changes, sleep disturbances, or other.

Reviewers recorded the overall attitude toward statins as perceived from the video (positive, negative, or neutral) and identified any potential false or misleading claims. Discrepancies between reviewers were resolved through discussion, with adjudication by a third reviewer when necessary. Potential claims were then independently reviewed by two subject matter experts, both cardiology fellows, and classified as true or false. Claims that were partially true, misleading, or lacking important clinical context were classified as false for the purposes of analysis. Subject matter expert disagreements were resolved by consensus. A video was considered to contain false claims if it included at least one false or misleading claim. True claims were excluded from subsequent analysis, whereas false claims were retained, counted, and grouped into meaningful content categories. Because videos could contain multiple false claims, the total number of false claims exceeded the number of videos with false claims.

### 2.4. Quality Assessment

Each video was independently assessed by two reviewers who were blinded to each other’s scores. Video quality was evaluated using the DISCERN instrument, the Patient Education Materials Assessment Tool for Audiovisual Materials (PEMAT-AV), and the Global Quality Score (GQS). These instruments were selected to assess multiple dimensions of video quality including reliability, understandability, actionability, and overall quality. All three tools have been widely used to evaluate online health information, including social media content [36–39].

The DISCERN instrument is a validated 16-item questionnaire designed to assess the quality and reliability of consumer health information related to treatment choices. The first section evaluates reliability, including clarity of aims, relevance, transparency of sources, balance, and discussion of uncertainties. The second section assesses the quality of information about treatment options, including benefits, risks, and support for shared decision-making. Each item is rated on a 5-point scale, with higher scores indicating higher quality [40]. A total DISCERN score was calculated for each video.

Understandability and actionability were assessed using the PEMAT-AV. This tool evaluates whether audiovisual materials are easy to understand and whether they provide clear, actionable steps for viewers. The Understandability domain assesses content clarity, word choice, organization, layout, and use of visual aids. The Actionability domain evaluates whether the material clearly identifies actions viewers can take and provides concrete, manageable steps. Items were scored as “Yes,” “No,” or “Not Applicable,” and percentage scores were calculated for each domain by dividing the number of “Yes” responses by the total number of applicable items [41].

Overall video quality was additionally assessed using the Global Quality Score (GQS), a 5-point Likert scale that evaluates overall educational value, flow, and usefulness to patients. Higher GQS scores indicate greater overall quality and usefulness [42]. For data analysis, scores from the two reviewers were averaged for each instrument.

### 2.5 Data Analysis

All analyses were performed using IBM SPSS Statistics version 31.0.0.0, with statistical significance set at p < 0.05. Categorical variables were summarized as frequencies and percentages. Continuous engagement variables were described using means, standard deviations, 95% confidence intervals, and medians.

Inter-rater reliability for DISCERN, PEMAT-AV, and GQS scores was evaluated using paired t-tests to assess systematic differences between reviewers, Pearson correlation coefficients to assess linear agreement, and intraclass correlation coefficients (ICC[2,1], two-way random effects, absolute agreement) to assess overall reliability.

Differences in quality scores across creator types were examined using one-way analysis of variance. Because homogeneity of variance was violated and group sizes were unequal, Welch’s ANOVA was used. Effect sizes were reported as eta-squared (η²). Given the larger effect size observed for DISCERN, post hoc pairwise comparisons were conducted for DISCERN scores using the Games–Howell procedure.

Differences in the prevalence of false claims across creator types were assessed using Pearson’s chi-square tests, with Cramer’s V reported as the measure of effect size. For creator-type analyses, false claims were summarized both as the number and percentage of videos within each creator group and as the proportion of all false-claim videos attributable to each creator group. False claims were also grouped into recurring content categories based on their subject matter. These categories were quantified using the total number of false claims as the denominator. Because individual videos could contain multiple false claims, category counts reflect the total number of false claims rather than the number of videos.

Associations between video views and quality scores were assessed using linear regression. Because view counts were positively skewed, log-transformed views were used where appropriate. Coefficients of determination (R²) were calculated to quantify the proportion of variance in quality scores explained by view count.

## 3. Results

Of the 1,349 original videos, 1,091 were excluded after manual review, for several reasons. For example, the term “Lipitor” generated Romanian-language videos referencing “lipitori” (leeches), “Crestor” returned platform-generated “creator insights” analytics videos, and “Mevacor” produced automobile-related content. Additionally, videos discussing cholesterol or lipid management without explicit reference to statins were excluded.

A total of 258 videos met inclusion criteria and were included in the final analysis. General statin-related terms (“statin,” “statins,” and “statin medication”) accounted for 44.6% of included videos. Among specific medications, atorvastatin (9.7%) and simvastatin (8.5%) were most frequently represented. Brand-specific searches yielded relatively few included videos, including Zocor (1.9%), Crestor (0.8%), and Pravachol (0.8%).

Creator types were diverse, with non-physician healthcare providers (34.5%) and physicians (26.0%) comprising the largest groups, followed by laypersons (18.2%), wellness promoters (12.8%), and complementary and alternative medicine professionals (8.5%). Most videos were categorized as educational (91.0%), with relatively few anecdotal or entertainment-focused posts.

Videos covered diverse themes related to the risks, benefits, and mechanisms of statin therapy. Negative effects were discussed more frequently (63.2%) than benefits (42.2%) or mechanism of action (30.6%). Among adverse effects, muscle pain or weakness was the most commonly mentioned (45.0%), followed by cognitive changes (22.1%) and metabolic or diabetes risk (19.4%). Among benefits, cholesterol lowering was most frequently cited (46.5%), followed by heart health (35.3%) and stroke prevention (23.3%), whereas longevity was referenced less often (5.8%). Overall, 44.2% of videos conveyed a neutral attitude toward statins, 32.6% were negative, and 23.3% were positive. False claims were identified in 39.5% of videos. See Table 1 for a summary of creator qualifications and video content.

**Table 1:**
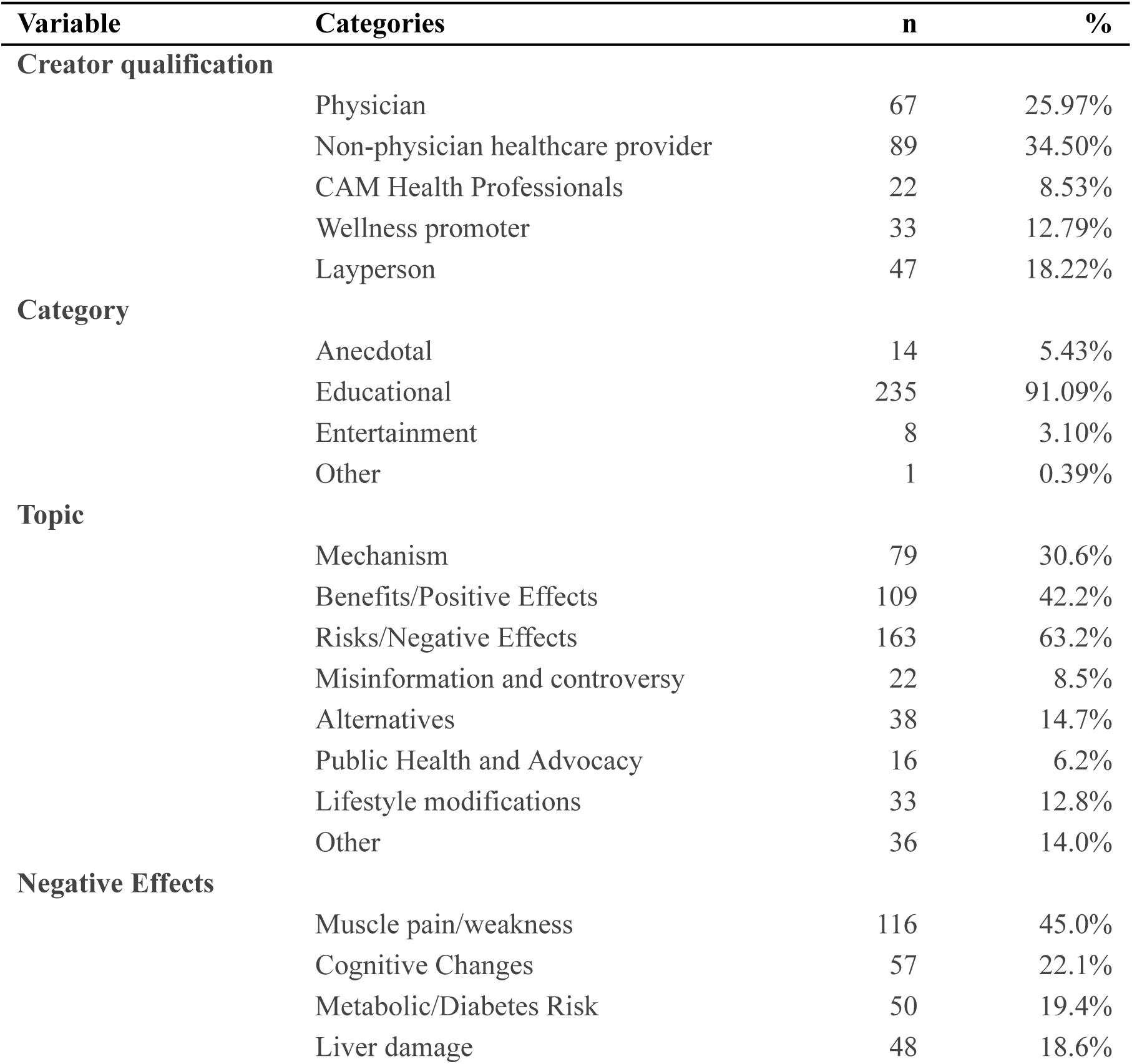
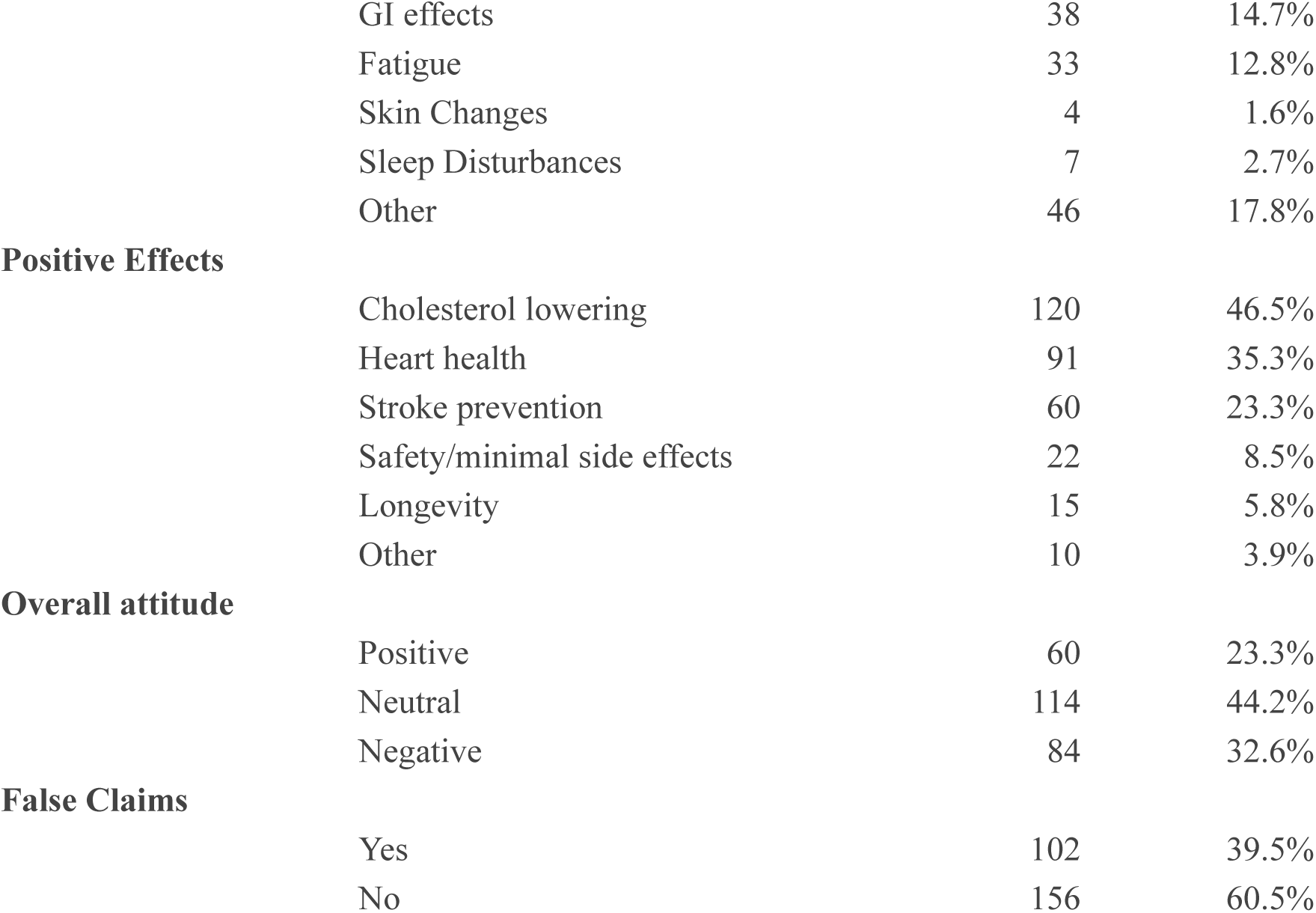
Creator qualifications and Video Content. Distribution of creator types, content categories, thematic topics, reported positive and negative effects, overall attitude toward statins, and prevalence of false claims among included videos (N = 258). Values are presented as frequencies and percentages. Abbreviations: CAM, complementary and alternative medicine

| Variable | Categories | n | % |
| --- | --- | --- | --- |
| **Creator qualification** |  |  |  |
|  | Physician | 67 | 25.97% |
|  | Non-physician healthcare provider | 89 | 34.50% |
|  | CAM Health Professionals | 22 | 8.53% |
|  | Wellness promoter | 33 | 12.79% |
|  | Layperson | 47 | 18.22% |
| **Category** |  |  |  |
|  | Anecdotal | 14 | 5.43% |
|  | Educational | 235 | 91.09% |
|  | Entertainment | 8 | 3.10% |
|  | Other | 1 | 0.39% |
| **Topic** |  |  |  |
|  | Mechanism | 79 | 30.6% |
|  | Benefits/Positive Effects | 109 | 42.2% |
|  | Risks/Negative Effects | 163 | 63.2% |
|  | Misinformation and controversy | 22 | 8.5% |
|  | Alternatives | 38 | 14.7% |
|  | Public Health and Advocacy | 16 | 6.2% |
|  | Lifestyle modifications | 33 | 12.8% |
|  | Other | 36 | 14.0% |
| **Negative Effects** |  |  |  |
|  | Muscle pain/weakness | 116 | 45.0% |
|  | Cognitive Changes | 57 | 22.1% |
|  | Metabolic/Diabetes Risk | 50 | 19.4% |
|  | Liver damage | 48 | 18.6% |
|  | GI effects | 38 | 14.7% |
|  | Fatigue | 33 | 12.8% |
|  | Skin Changes | 4 | 1.6% |
|  | Sleep Disturbances | 7 | 2.7% |
|  | Other | 46 | 17.8% |
| **Positive Effects** |  |  |  |
|  | Cholesterol lowering | 120 | 46.5% |
|  | Heart health | 91 | 35.3% |
|  | Stroke prevention | 60 | 23.3% |
|  | Safety/minimal side effects | 22 | 8.5% |
|  | Longevity | 15 | 5.8% |
|  | Other | 10 | 3.9% |
| **Overall attitude** |  |  |  |
|  | Positive | 60 | 23.3% |
|  | Neutral | 114 | 44.2% |
|  | Negative | 84 | 32.6% |
| **False Claims** |  |  |  |
|  | Yes | 102 | 39.5% |
|  | No | 156 | 60.5% |

Video length and engagement metrics demonstrated substantial variability (Table 2). Videos had a mean duration of 92.3 seconds (SD 96.0), with a median of 64.5 seconds (IQR, 41.0–106.5). View counts ranged from 101 to 2,100,000 and were positively skewed, with a mean of 90,074 (SD 247,263) and a substantially lower median of 16,850 (IQR, 4,561–55,050). Similar right-skewed distributions were observed for comments, shares, and saves, with mean values consistently exceeding medians. The mean number of comments was 142 (SD 311), shares 1,029 (SD 4,394), and saves 832 (SD 3,096). The mean engagement rate was 3.61% (SD 2.47%), with a median of 3.17% (IQR, 1.84%–4.73%).

**Table 2:** Video Metrics. Descriptive statistics for video length and engagement metrics, including views, comments, shares, saves, and engagement rate (sum of likes, comments, shares, and saves divided by total views). Continuous variables are presented as minimum, maximum, mean (standard deviation), 95% confidence interval (lower and upper confidence limits), and median (interquartile range). Abbreviations: SD, standard deviation; CI, confidence interval; LCL, lower confidence limit; UCL, upper confidence limit.

| Video characteristic | Minimum | Maximum | Mean (SD) | 95% CI of mean (LCL, UCL) | Median (Q1, Q3) |
| --- | --- | --- | --- | --- | --- |
| **Video length (s)** | 5 | 573 | 92.3 (96.0) | (80.5, 104.0) | 64.5 (41.0, 106.5) |
| **Likes** | 2 | 89,500 | 2276 (8578) | (1124, 3327) | 231 (88, 885) |
| **Views** | 101 | 2,100,000 | 90,074 (247,263) | (59,760, 120,388) | 16,850 (4,561–55,050) |
| **Comments** | 0 | 2,124 | 142 (311) | (103, 180) | 28 (7, 129) |
| **Shares** | 0 | 42,700 | 1,029 (4,394) | (490, 1,568) | 42 (8, 159) |
| **Saves** | 0 | 29,200 | 832 (3,096) | (452, 1,211) | 51 (16, 221) |
| **Engagement rate** | 3.31% | 3.91% | 3.61% (2.47%) | (3.31%, 3.91%) | 3.17% (1.84%, 4.73%) |

Mean quality scores were generally in the low-to-moderate range across instruments (Table 3). Inter-rater reliability was strong overall. No statistically significant differences were observed between reviewers for DISCERN (mean difference 0.15, p = 0.684) or PEMAT–Understandability (mean difference 0.2%, p = 0.761). Although small but statistically significant differences were observed for PEMAT–Actionability (mean difference −3.6%, p = 0.002) and Global Quality Scale (mean difference 0.16, p = 0.001), the magnitude of these differences was small relative to the overall scale ranges. Pearson correlation coefficients demonstrated high agreement between reviewers across all measures (r = 0.814–0.907, all p < 0.001). Intraclass correlation coefficients indicated good to excellent reliability, ranging from 0.808 (GQS) to 0.906 (DISCERN). Additionally, the majority of ratings fell within predefined agreement thresholds across instruments (69.0%–93.8%).

**Table 3:** Video Quality and Inter-Rater Reliability. Inter-rater comparison of DISCERN, PEMAT–AV (Understandability and Actionability), and Global Quality Scale (GQS) scores. Values include reviewer means (SD), mean differences with 95% confidence intervals, paired t-test results, Pearson correlation coefficients, intraclass correlation coefficients (ICC; two-way random effects, absolute agreement), and predefined agreement thresholds. Abbreviations: DISCERN, Quality of Consumer Health Information Instrument; PEMAT–AV, Patient Education Materials Assessment Tool for Audiovisual Materials; GQS, Global Quality Scale; ICC, intraclass correlation coefficient. REV, Reviewer; CI, Confidence Interval.

| Measure | REV 1<br>Mean<br>(SD) | REV 2<br>Mean<br>(SD) | Mean<br>Difference<br>(95% CI) | Paired<br>t-test p<br>value | Pearson r<br>(p value) | ICC (95% CI) | Within Threshold<br>n (%) |
| --- | --- | --- | --- | --- | --- | --- | --- |
| DISCERN | 38.2<br>(14.1) | 38.1<br>(13.3) | 0.15 (−0.58<br>to 0.88) | 0.684 | 0.907<br>($<0.001$) | 0.906<br>(0.882–0.926) | Within 5 points:<br>178 (69.0%) |
| PEMAT –<br>Understandability | 63.0%<br>(21.0%) | 63.0%<br>(20.0%) | 0.2% (−1.2%<br>to 1.6%) | 0.761 | 0.848<br>($<0.001$) | 0.848<br>(0.810–0.879) | Within 10% (0.10):<br>224 (86.8%) |
| PEMAT –<br>Actionability | 46.0%<br>(38.0%) | 50.0%<br>(38.0%) | −3.6%<br>(−6.0% to<br>−1.3%) | 0.002 | 0.877<br>($<0.001$) | 0.874<br>(0.839–0.901) | Within 10% (0.10):<br>189 (73.3%) |
| Global Quality<br>Scale (GQS) | 2.77<br>(1.30) | 2.93<br>(1.31) | 0.16 (−0.26<br>to −0.06) | 0.001 | 0.814<br>($<0.001$) | 0.808<br>(0.758–0.848) | Within 1 point: 242<br>(93.8%) |

Quality scores differed significantly across creator types for all assessment instruments (Table 4). Welch’s ANOVA demonstrated statistically significant differences in DISCERN total scores (Welch F(4, 95.98) = 40.22, p < 0.001, η² = 0.341), PEMAT–Understandability (Welch F(4, 88.67) = 11.27, p < 0.001, η² = 0.143), PEMAT–Actionability (Welch F(4, 87.74) = 5.60, p < 0.001, η² = 0.079), and Global Quality Scale scores (Welch F(4, 253) = 27.07, p < 0.001, η² = 0.300). Physicians consistently demonstrated the highest mean scores across all measures, including DISCERN (48.19 [SD 11.87]), PEMAT–Understandability (72.5% [SD 15.2%]), PEMAT–Actionability (61.1% [SD 35.1%]), and GQS (3.69 [SD 0.98]). In contrast, complementary and alternative medicine providers, wellness promoters, and laypersons exhibited lower mean scores, particularly for DISCERN and GQS.

**Table 4.**
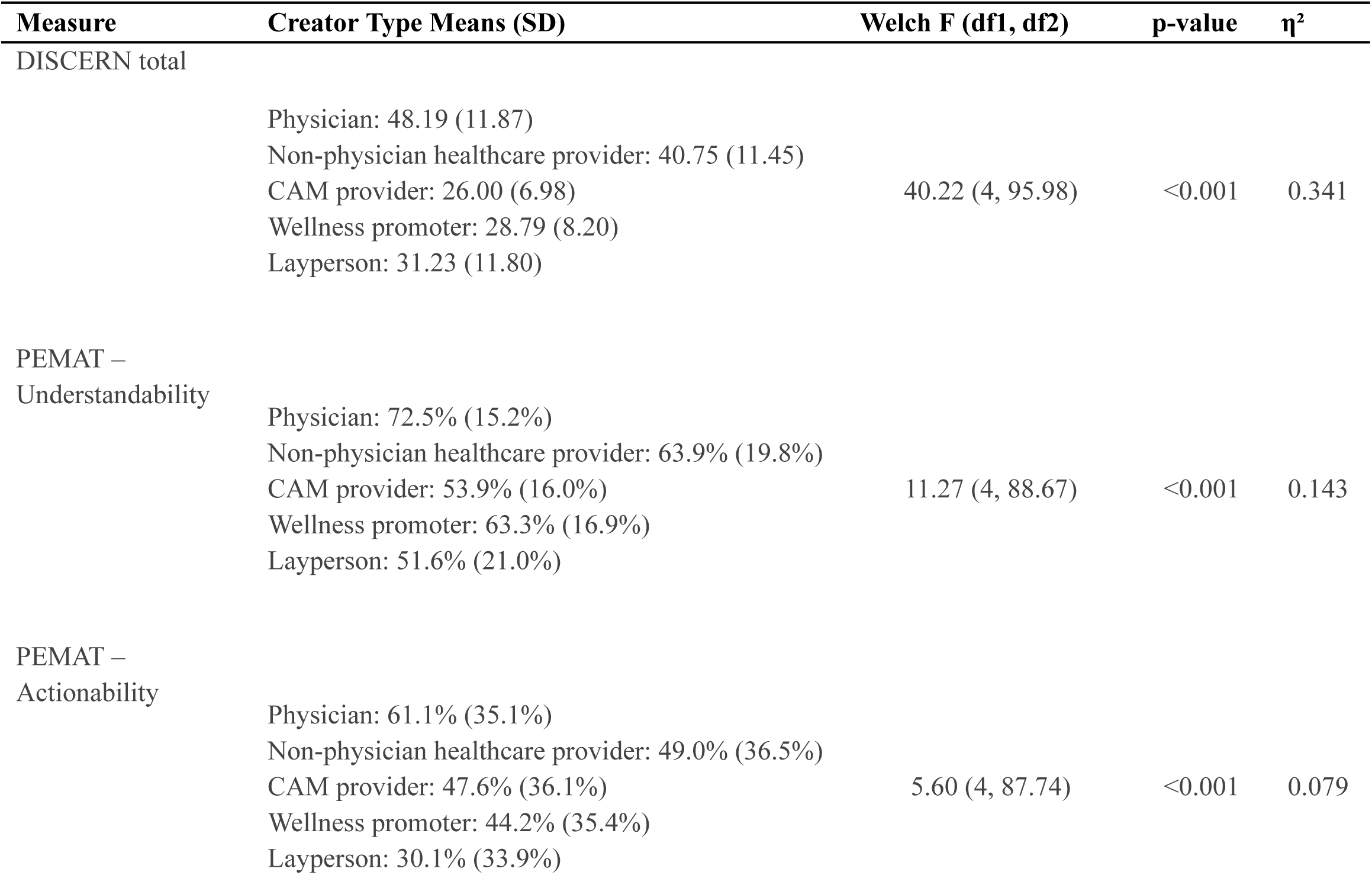
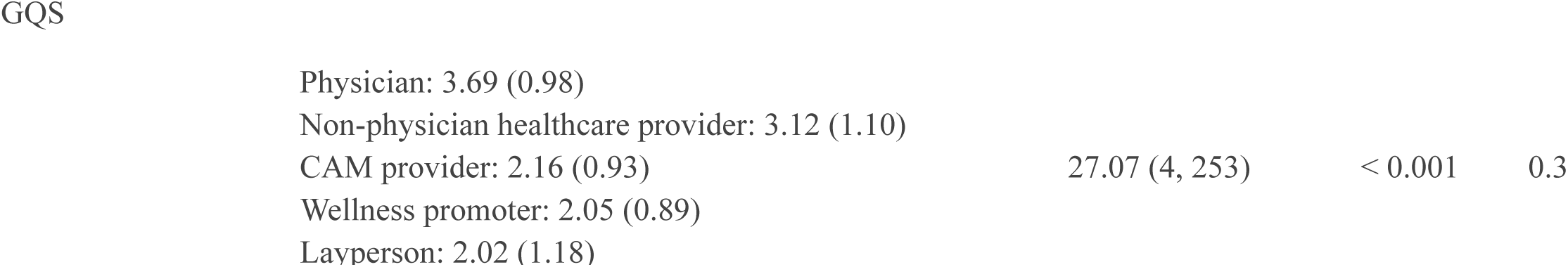
Differences in Quality Scores Across Creator Types. Comparison of mean quality scores across creator types using Welch’s ANOVA to account for unequal variances and group sizes. Values represent mean (SD) for each creator group. Welch F statistics, p values, and eta-squared (η²) effect sizes are reported. Post hoc Games–Howell comparisons for DISCERN demonstrated Physicians > all groups (mean differences 7.43–22.19, all p ≤ 0.001); Non-physician healthcare providers > CAM, wellness promoters, and laypersons (mean differences 9.52–14.75, all p < 0.001); CAM, wellness promoters, and laypersons did not differ significantly (all p > 0.05). Abbreviations: SD, standard deviation; DISCERN, Quality of Consumer Health Information Instrument; CAM, complementary and alternative medicine; η², eta-squared; PEMAT–AV, Patient Education Materials Assessment Tool for Audiovisual Materials; GQS, Global Quality Scale.

Effect sizes indicated that creator type explained a substantial proportion of variance in DISCERN (η² = 0.341) and GQS (η² = 0.300), and a more modest proportion of variance in PEMAT–Understandability (η² = 0.143) and PEMAT–Actionability (η² = 0.079). Post hoc Games–Howell comparisons for DISCERN total scores demonstrated that physicians scored significantly higher than all other creator groups (all p ≤ 0.001). Non-physician healthcare providers also scored significantly higher than complementary and alternative medicine providers, wellness promoters, and laypersons (all p < 0.001). No statistically significant differences were observed among complementary and alternative medicine providers, wellness promoters, and laypersons.

False claims varied significantly by creator type (Pearson χ²(4, N = 258) = 66.92, p < 0.001; Cramér’s V = 0.51) (Table 5). Although wellness promoters accounted for the largest share of all false-claim videos (26.5%), the highest within-group prevalence was observed among CAM health professionals (90.9%), followed by wellness promoters (81.8%). In contrast, physicians and non-physician healthcare providers had the lowest within-group prevalence of false claims, at 25.4% and 21.3%, respectively. They also accounted for smaller shares of the total false-claim videos, with physicians contributing 16.7% and non-physician healthcare providers contributing 18.6% of all false-claim content.

**Table 5.** Prevalence of False Claims by Creator Type. Presents the number and proportion of videos containing at least one false claim across creator types, including both the percentage within each creator group and the percentage of total false-claim videos contributed by each group. The table also reports the Pearson chi-square test and Cramer’s V effect size for differences in false-claim prevalence by creator type. Abbreviations: CAM, complementary and alternative medicine; Pearson χ², Pearson chi-square.

| Creator Type | False Claims n | % within creator type | % of total false claims |
| --- | --- | --- | --- |
| Physician | 17 | 25.40% | 16.67% |
| Non-physician healthcare provider | 19 | 21.30% | 18.63% |
| CAM health professionals | 20 | 90.90% | 19.61% |
| Wellness promoter | 27 | 81.80% | 26.47% |
| Layperson | 19 | 40.40% | 18.63% |
| Total | 102 | 39.50% | 100.00% |

Pearson $\chi^2(4, N = 258) = 66.92, p < 0.001$

False claims encompassed a broad range of themes (Table 6). The most frequently identified categories involved alleged coenzyme Q10 depletion (n = 26, 11.6% of coded false claims), claims that statins reduce cholesterol needed for hormone production (n = 18, 8.0%), promotion of alternatives to statin therapy (n = 14, 6.3%), and denial of the causal role of low-density lipoprotein cholesterol in heart disease (n = 14, 6.3%). An additional 82 claims (36.6%) were categorized as miscellaneous because they did not fall within a broader recurring theme. Examples of miscellaneous themes include claims that most patients experience quality-of-life-limiting side effects (n = 1, 0.45%) from statins and that statins can cause migraines (n = 1, 0.45%).

**Table 6.** Categories of False Claims About Statins. Summarizes the coding framework used to classify 224 false-claim instances about statins in the dataset. It lists each false claim category, the count of instances in that category (n), and the percentage of the total sample (%). Abbreviations: CoQ10, coenzyme Q10; LDL/LDLs, low-density lipoprotein(s).

| **False claim category** | **n** | **%** |
| --- | --- | --- |
| Statins significantly lower CoQ10 levels. | 26 | 11.61% |
| Statins reduce cholesterol needed for hormone production. | 18 | 8.04% |
| Alternatives | 14 | 6.25% |
| Increased levels of LDLs don't cause heart disease. | 14 | 6.25% |
| Statins increase risk of dementia. | 13 | 5.80% |
| High levels of cholesterol is not bad for your health. | 13 | 5.80% |
| Statins cause significant muscle damage. | 9 | 4.02% |
| Statins don't prevent heart attack or stroke. | 9 | 4.02% |
| Statins reduce cholesterol in the brain. | 8 | 3.57% |
| Statins cause significant liver damage. | 7 | 3.13% |
| Statins significantly deplete vitamin D levels. | 6 | 2.68% |
| Statins cause mood changes or depression. | 5 | 2.23% |
| Miscellaneous | 82 | 36.61% |
| **Total** | **224** | **100.00%** |

Linear regression analyses demonstrated minimal association between video views and quality scores across all instruments (Figure 1). Although slight positive slopes were observed, coefficients of determination were low (R² range, 0.001–0.033), indicating that view count explained between 0.1% and 3.3% of the variance in DISCERN, PEMAT–Understandability, PEMAT–Actionability, and GQS scores. Even for PEMAT–Actionability, which demonstrated the largest R² (0.033), the magnitude of association remained small. Visual inspection of scatterplots showed wide dispersion of quality scores across all levels of video popularity.

**Figure 1:**
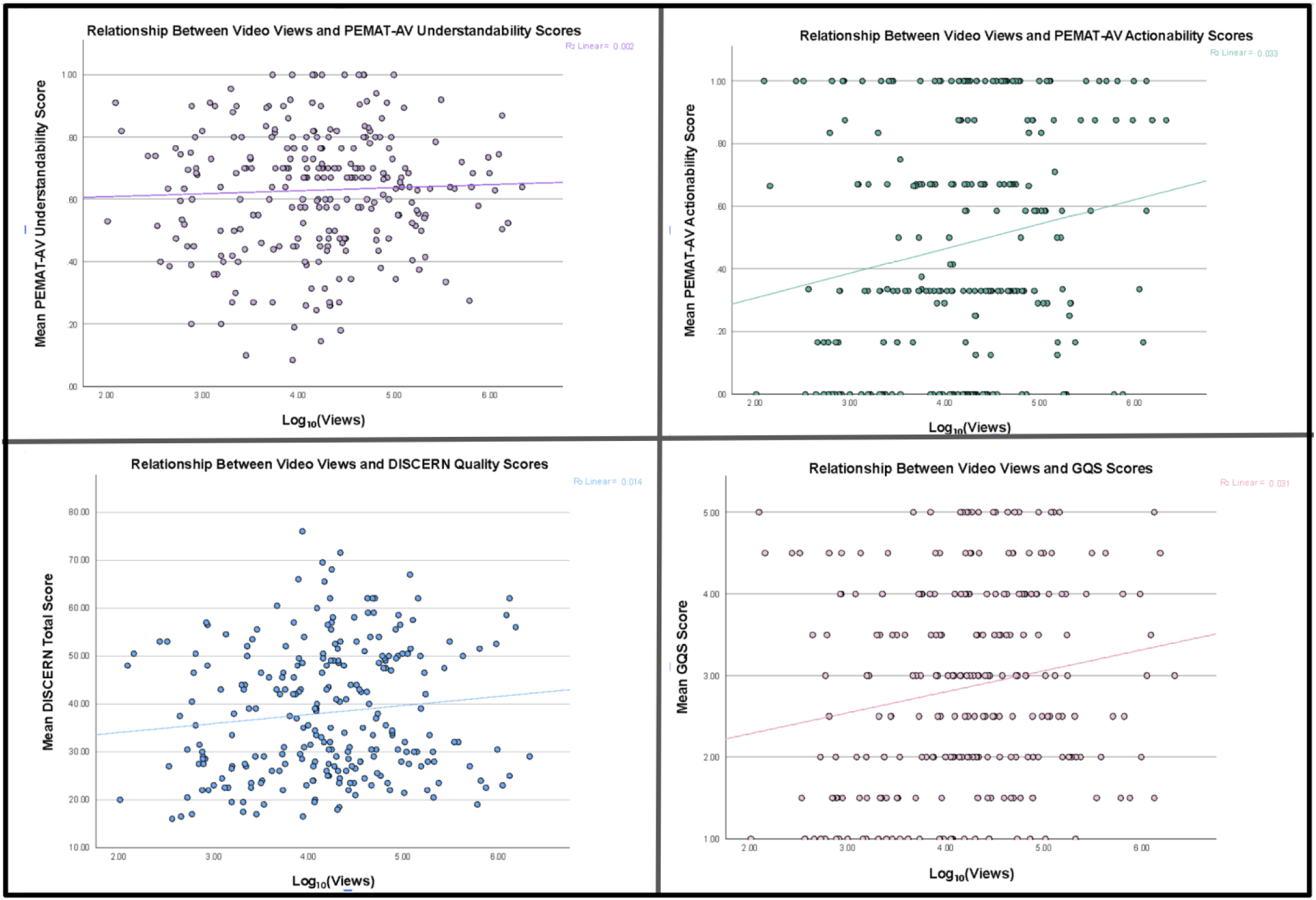
Relationship Between Log-Transformed Video Views and Quality Scores. Scatterplots depicting the relationship between log₁₀-transformed video views and DISCERN total scores, PEMAT–AV Understandability scores, PEMAT–AV Actionability scores, and Global Quality Scale (GQS) scores. Each point represents an individual video. Solid lines represent fitted linear regression models. Coefficients of determination (R²) are displayed for each panel. Abbreviations: DISCERN, Quality of Consumer Health Information Instrument; PEMAT–AV, Patient Education Materials Assessment Tool for Audiovisual Materials; GQS, Global Quality Scale.

## 4. Discussion

### 4.1 Overview of Results

This study provides a comprehensive evaluation of statin-related content on TikTok and reveals several important patterns regarding information quality, misinformation, and engagement. Thematic analysis revealed that potential risks and negative effects of statins were discussed more frequently than benefits, appearing in 63.2% of videos compared with 42.2% that discussed positive effects. Among the negative effects mentioned, muscle pain or weakness was the most common, followed by cognitive changes and metabolic risk, while the most frequently cited benefits included cholesterol lowering, heart health, and stroke prevention. Although overall sentiment toward statins was most often neutral, a substantial proportion of videos conveyed negative attitudes. Together, these findings indicate that TikTok discussions of statins may emphasize potential harms more prominently than benefits, which may influence how viewers perceive the role of statin therapy in cardiovascular prevention.

This thematic imbalance mirrors patterns observed in prior statin-specific social media studies on text-based platforms. Golder and colleagues [11] found that content regarding adverse effects were the most prominent in statin-related Twitter (now known as X) posts. Similarly, Somani and colleagues (2023) identified adverse effects, hesitancy, pharmaceutical industry bias, and supplement or lifestyle alternatives as dominant Reddit themes, with discussions showing predominantly neutral-to-negative sentiment [16]. Slavin and colleagues [43] further demonstrated that negative sentiment and statin skepticism on Twitter/X increased over time, and negative sentiment was associated with greater engagement.

The greater emphasis on risks over benefits may be a reflection of both psychological and commercial influences. Negative information tends to attract more attention and carry greater weight than positive information, a pattern often described as negativity bias [44,45]. Content that highlights side effects, controversy, or mistrust may therefore be more likely to attract engagement and be amplified through platform algorithms. In addition, creators who promote supplements, alternative therapies, or wellness products may benefit from framing statins negatively, as this framing can direct viewers toward alternative treatments or services. The short-form structure of TikTok videos may further reinforce this imbalance by encouraging simplified or emotionally engaging narratives rather than nuanced discussions of risk–benefit tradeoffs.

Our analysis also demonstrated that video quality differed significantly by creator type, with physicians consistently producing the highest-quality content across all assessment instruments (DISCERN, PEMAT–Understandability, PEMAT–Actionability, and Global Quality Score). In contrast, videos produced by CAM providers, wellness promoters, and laypersons demonstrated substantially lower quality scores. These differences were not only statistically significant but also meaningful in magnitude, with creator type accounting for a large proportion of the variation in DISCERN and GQS scores. Post hoc analysis comparisons further reinforced these findings by showing that physician-created content scored significantly higher than all other creator groups, while non-physician healthcare providers also outperformed CAM creators, wellness promoters, and laypersons. In contrast, the lack of significant differences among CAM creators, wellness promoters, and laypersons suggests that these groups produced statin content of similarly low quality, despite differences in how their expertise was presented.

These findings are consistent with previous social media studies showing that physicians or other professionals with healthcare degrees tend to create higher quality and more reliable content. As expected, TikTok studies in a wide variety of medical topics all demonstrate that healthcare professionals created videos scored higher on the DISCERN, GQS, and PEMAT-AV, although these videos did not necessarily receive higher rates of engagement than content created by non-healthcare influencers [20, 21, 46–48]. Our findings reinforce this pattern and highlight the persistent gap between informational quality and online visibility.

The observed differences in quality likely reflect differences in training and familiarity with evidence-based medicine. Physicians receive formal training in interpreting clinical evidence, evaluating treatment risks and benefits, and communicating complex medical information. This training may enable them to present statin therapy in a more balanced and accurate manner, which aligns with the criteria evaluated by instruments such as DISCERN and GQS. By comparison, nonmedical creators may rely more heavily on audience friendly framing and personal perspectives that feel relatable to viewers. Although such approaches may increase relatability and engagement, they may also omit important clinical nuance or present incomplete interpretations of the evidence. Differences across creator types were somewhat smaller for PEMAT-Understandability and Actionability because these tools focus more on how the information is presented than the content as a whole. As a result, creators without formal medical training may still produce content that appears easy to understand or engaging to viewers, even when the underlying information is incomplete, misleading, or poorly supported.

Our results also revealed that highly viewed videos were not necessarily more accurate or informative than less viewed content. View counts explained only a very small proportion of the variance in DISCERN, PEMAT-AV, and GQS scores, indicating that video popularity was a poor predictor of informational quality. Both high- and low-quality videos appeared across all levels of popularity. These results are similar to findings from other social media studies. For example, Zeng and colleagues [24] found no significant association between engagement and accuracy or evidence level in nutrition content on TikTok, while Shackleford and colleagues [20] reported that oral-contraceptive videos from healthcare professionals were more reliable yet received fewer views, likes, and comments than videos from non-healthcare creators.

The lack of association between views and quality likely reflects the dynamics of TikTok’s algorithm-driven recommendation system, in which content visibility is influenced primarily by engagement signals, such as watch behavior, shares, and other interaction patterns, rather than informational accuracy [23]. As a result, content that is brief, emotionally compelling, humorous, relatable, or controversial may therefore achieve greater visibility regardless of its educational value. Perceived popularity may also influence viewer trust, as highly viewed videos can appear more credible simply because they seem widely endorsed. These dynamics create an environment in which both high and low quality videos can achieve substantial reach, making engagement metrics a poor indicator of informational value.

Another important finding of this study was the high prevalence of false or misleading claims with nearly 40% of videos containing at least one inaccurate statement. False claims were strongly associated with creator type and were not distributed evenly across groups and were concentrated disproportionately among CAM health professionals and wellness promoters, while physician and non-physician healthcare provider accounts demonstrated substantially lower rates of false claims. The content of these claims was particularly concerning as many challenged established foundational concepts in lipid biology and cardiovascular prevention. For example, claims denying a causal role for low-density lipoprotein cholesterol in heart disease contradict extensive genetic, epidemiologic, and clinical trial evidence showing that LDL is a causal driver of atherosclerotic cardiovascular disease and that LDL lowering reduces cardiovascular events [2,49].

Another common false claim was that statins “deplete” coenzyme Q10 in a way that causes fatigue, cognitive symptoms, or statin-induced myopathy. Although randomized trials and meta-analyses show that statin therapy can lower circulating plasma CoQ10 levels, this appears to largely track with reductions in LDL-containing lipoprotein carriers rather than proving clinically meaningful tissue depletion. More recent muscle-based data have found unaltered intramuscular CoQ10 levels despite statin exposure [50,51,53]. These claims may be especially persuasive because they present nonpharmacologic options as safer or more “natural,” potentially discouraging use of therapies with proven cardiovascular benefit. Statins are long-term preventive therapies, and public perceptions of their risks and benefits can directly influence acceptance, adherence, and continuation. When combined with our finding that risks and negative effects were discussed more often than benefits, these results suggest that TikTok may expose users to a skewed representation of statin therapy that overemphasizes harms while underrepresenting established benefits.

The prevalence and nature of these claims are consistent with findings from other social media studies evaluating therapeutic misinformation. For example, in TikTok studies about sinusitis, less than half of the videos posted by nonmedical influencers contained accurate information [21]. In nutrition-focused Tiktok research, 75% of videos lacked balanced and accurate content [24]. These studies similarly found that nonmedical creators frequently disseminated inaccurate or misleading health information, particularly when promoting alternative therapies or wellness products.

The high prevalence of false claims in our sample may reflect the inherent tension between the complexity of statin therapy and the simplicity favored by short form social media. Accurate discussion of statins often requires nuanced explanation of risk reduction, side effects, patient selection, and long term prevention. However, TikTok content is typically brief and optimized for quick, memorable takeaways [54]. In that setting, nuanced evidence may be compressed into oversimplified statements that are easier to understand but more likely to become misleading. As with the predominance of negative sentiment toward statins across social media platforms, false claims may be more likely to arise because statins are a controversial and emotionally charged topic, making them particularly vulnerable to selective framing, exaggerated harms, and repeated misconceptions. When viewed together these factors may help explain why false or misleading claims were so common in our sample.

### 4.2 Implications

These findings have several important implications for the evolving role of social media in healthcare. This study demonstrated that engagement metrics were not associated with informational quality; therefore, users cannot reliably use popularity as a proxy for credibility when evaluating statin-related content. This dynamic creates a challenging environment in which inaccurate or misleading information may achieve substantial visibility and influence public perceptions of widely used preventive therapies. At the same time, the widespread reach of social media platforms presents an opportunity for healthcare workers to engage with the community and a broader audience than they would otherwise encounter in a clinical setting, and disseminate evidence-based medical information. Proactive social media engagement by cardiologists, preventative medicine specialists, and professional societies presents a vital opportunity to advance public health education. As an accessible and relevant platform, social media offers an ideal channel to reach many patients that may not have regular healthcare follow-up.

The dissemination of medically inaccurate or misleading information could further exacerbate the existing issue of health literacy. Without credible sources to counterbalance inaccurate and low informational quality videos, there is further discrepancy in communication between physicians and patients. The themes identified in this study also provide insight into common concerns patients have regarding statins and offer a valuable insight into concerns circulating in online discussions of statins. Awareness of these concerns may help clinicians address misconceptions more effectively during patient counseling and support informed decision-making to promote statin adherence and therefore prevention of major cardiovascular events further improving patient care.

### 4.3 Strengths and Limitations

This study has several strengths. To our knowledge, it is the first study to specifically examine statin-related content on TikTok, and one of the first to systematically evaluate the quality of statin-related social media content using multiple validated quality instruments simultaneously, including DISCERN, PEMAT-AV, and GQS. By applying these complementary tools, the study was able to provide a multidimensional assessment of informational quality, reliability, understandability, and actionability. In addition, this study is among the first to quantify the prevalence of false or misleading claims related to statins on TikTok and to characterize the types of misinformation being shared. By doing so, our study extends prior statin research by providing a clearer understanding of the misinformation users may encounter about statins on social media, and which creator group may be contributing most to that misinformation.

The systematic search and defined inclusion criteria provided a comprehensive evaluation of the statin-related content available to users on the platform. The relatively large sample of 258 videos likely reflects the content most commonly encountered by individuals searching for statin-related information on the platform. In addition, employing several established instruments allowed for a multidimensional assessment of video quality, reliability, understandability, and actionability, which strengthened the overall evaluation of health information quality.

Another important strength of the study is the rigorous evaluation of inter-rater reliability between reviewers which enhanced the internal consistency of the scoring process and increased confidence in the objectivity of the assessment, reflecting true characteristics of the videos rather than chance variation or reviewer subjectivity. This rigor was further strengthened by independent review of potential false claims by two subject matter experts. Each claim was evaluated against reputable peer-reviewed evidence and any disagreements were resolved through active discussion and critical examination of one another’s interpretations. In addition, the application of robust statistical modeling to analyze associations between video characteristics, quality metrics, and engagement further strengthened the findings of the study.

Several limitations should be considered. First, the cross-sectional design captures content at a single point in time and may not reflect the rapidly evolving nature of TikTok content or algorithmic recommendations. Additionally, videos originally identified during the search process were also unavailable at the time of analysis. Video availability, engagement metrics, and platform algorithms change rapidly, resulting in findings that may represent a snapshot rather than a permanent characterization of available content.

Second, the study focused exclusively on TikTok as a platform for video collection limiting the generalizability of the study to other social media platforms such as Instagram, Facebook, etc. This study may not capture the overall public sentiment toward statins, but rather TikTok users’ views of statins. Restricting the analysis to English-language videos also limits generalizability to non–English-speaking audiences.

Finally, although DISCERN, PEMAT-AV, and GQS are widely used and valid tools of measurement, they were developed to analyze more traditional longer-form educational video content. Their application to short-form social media videos may have inherent limitations. Nevertheless, these tools remain among the most widely used and validated methods for evaluating online health information and allow for a standardized and systematic assessment of video quality. Despite these limitations, this study provides valuable insight into the quality of commonly consumed video content available to the general public and reveals opportunities for healthcare providers to become more involved in public health education via social media engagement.

### 4.4 Future Directions

This study adds to the growing body of evidence regarding the impact of social media on public health. Future research could further explore the complexities of how public health information is shared across these platforms. Because we used a cross-sectional design, we were unable to track statin-related misinformation over time. A longitudinal approach could offer valuable insight into the rapidly evolving public health landscape and how it interacts with current events. Additionally, examining multiple platforms would provide a more comprehensive view of the available information and may reveal differences in content trends. This would be particularly useful, as different social media platforms tend to attract distinct audience demographics.[55].

Our cross-sectional study did not allow us to assess the impact of any interventions. Future studies could explore the effects of experimentally amplifying expert-generated content and its potential influence on public health literacy. A closer look at how the general public perceives expert content creators may also help clarify how this information is received and offer additional insight into digital health literacy. Beyond perception, behavioral change is another important outcome to consider—for example, whether exposure to expert versus non-expert content affects health-related decisions.

Understanding how these videos influence viewer behavior would help identify potential interventions to improve digital health literacy and promote evidence-based medicine beyond the clinical setting

## 5. Conclusion

This study provides a comprehensive evaluation of statin-related content on TikTok and demonstrates a strong association between creator type and information quality. Physician-created videos are of higher quality and reliability than content produced by other creator groups. Notably, these differences in information quality were not reflected in measures of video popularity, as engagement metrics were similar across both low- and high-quality videos. False claims remained common, with the highest prevalence observed among CAM health professionals and wellness promoters and the lowest among physicians and non-physician healthcare providers. In addition, statin-related discussions on TikTok more often emphasized potential harms than established cardiovascular benefits. These findings underscore the need for increased participation by healthcare professionals in social media health communication and for improving digital health literacy in an environment where popularity does not necessarily reflect accuracy.

## Data Availability Statement

The data and coding framework supporting the findings of this study are available from the corresponding author upon reasonable request.

## CRediT Author Contributions

**Iren Gharibyan:** Conceptualization, Methodology, Investigation, Data curation, Formal analysis, Visualization, Writing – original draft, Writing – review and editing.

**Emily Ahner:** Conceptualization, Methodology, Investigation, Data curation, Validation, Writing – original draft, Writing – review and editing.

**Ryan Shao:** Validation, Writing – review and editing.

**Divyansh Sharma:** Validation, Writing – review and editing.

**Tracy Navarsartian Tazehkand:** Investigation, Data curation.

**Jimmy Diep:** Supervision, Validation, Writing – review and editing.

**Bertille Assoumou:** Conceptualization, Methodology, Supervision, Project administration, Writing – review and editing.

## Acknowledgements

The authors have no acknowledgements to declare.

